# Lutetium-177 hydroxyapatite radiosynovectomy in refractory chronic inflammatory arthritis of the knee joint

**DOI:** 10.1101/2024.10.06.24314955

**Authors:** Tanvi Sarwal, Rajesh Kumar, Sameer Taywade, Maya Gopalakrishnan, Abhay Elhence, Siyaram Didel, Ravi Gaur

## Abstract

**Objective:** To determine the efficacy of ^177^Lutetium hydroxyapatite radiosynovectomy (RSV) in patients with refractory chronic inflammatory arthritis of the knee joint.

**Materials and Methods:** Overall, 24 knees in 22 patients with refractory chronic inflammatory arthritis were enrolled in this prospective study. All patients were assessed clinically for pain, tenderness, range of motion, analgesic intake, Oxford knee score and blood pool activity on three phase bone scan. Various scores were assigned to clinical these parameters. RSV of the knee joint was done using intra-articular injection of ^177^Lutetium hydroxyapatite. Patients were assessed clinically at 1 and 3 months, and scintigraphically at 3 months using blood pool index on three phase bone scan. Patients were categorised as responders and non-responders on the basis of change in percentage of cumulative scores.

**Results:** Out of 24 knees, 18 knees were responders and 6 knees were non-responders at 3 months. There was a significant improvement in clinical scores at 1 month post RSV, which persisted at 3 months post RSV (p < 0.05). However, the change in blood pool activity on three phase bone scan was not significant. Reported adverse effects were mild and not significant.

**Conclusion:** This study confirmed the safety and efficacy of ^177^Lutetium hydroxyapatite radiosynovectomy for patients with chronic inflammatory knee joint arthritis refractory to conventional medical treatment.

## INTRODUCTION

Arthritis is an important cause of knee pain. The term is derived from the Greek term “arthos” meaning joint and “itis” meaning inflammation, thus arthritis is an inflammation of the joints (1). Broadly, arthritis can be divided into two main types: inflammatory and degenerative/non-inflammatory. The most common cause of inflammatory arthritis is rheumatoid arthritis, while osteoarthritis is the most common cause of non-inflammatory arthritis (2).

The symptoms that a patient experiences will vary depending on the type of arthritis they have. For example, in inflammatory arthritis, patients commonly present with pain, swelling, warmth, and tenderness in the involved joints (2). In most of the inflammatory arthritis, the main pathology is that of synovitis (3). Synovitis means inflammation of the synovium, the special connective tissue lining of a joint cavity (4). Although synovitis was classically associated with inflammatory disorders like rheumatoid arthritis, mounting evidence suggests that osteoarthritis also has an inflammatory component, and synovitis plays an important component in the pathogenesis of osteoarthritis as well (5, 6, 7).

Treatment options for inflammatory arthritis include symptomatic relief with non-steroidal anti-inflammatory drugs (NSAIDs) to manage pain and inflammation. Disease-modifying anti-rheumatic drugs (DMARDs) and biologics are prescribed to slow the progression of the disease (8). While many patients respond well to these treatments, some patients do not, and the relief is often temporary, with a high likelihood of recurrence after discontinuation. Additionally, DMARDs and biologics can cause serious side effects and are expensive, limiting their long-term use (8, 9).

In patients with refractory inflammatory arthritis, synovectomy is a potential treatment modality. Synovectomy can be carried out in 3 ways: surgical, chemical, and radiation synovectomy (10). Although surgical synovectomy is successful, it is costly, and prolonged rehabilitation is required. Chemical synovectomy involves the usage of chemicals like rifampicin or osmic acid. It is painful, and the long-term response is relatively low. Further, there is a risk of massive haemorrhage in hemophilia patients (11, 12).

Nuclear medicine has an attractive alternative option for treating refractory synovitis of joints, that is radiation synovectomy. The basic principle in radiation synovectomy is centred around destroying the inflamed synovium, which is producing inflammatory mediators (3). The procedure involves injecting a beta-emitting radionuclide into the joint cavity. The radionuclide is phagocytosed by the macrophages of the synovial membrane. The radionuclide emits beta rays, which lead to apoptosis of the synovial cells, and eventual fibrosis of the synovial membrane. Thus, synovial fluid production is decreased, and symptoms of joint inflammation reduce (13, 14).

A wide range of radionuclides are available for RSV worldwide. The European Association of Nuclear Medicine (EANM) guidelines recommend three radionuclides, namely ^90^Yttrium, ^186^Rhenium, and ^169^Erbium (15). Other radionuclides are also being used, like ^32^Phosphorous, ^165^Dysprosium, ^188^Rhenium, ^166^Holmium, ^153^Samarium and more recently, ^177^Lutetium (16).

^177^Lutetium is a relatively recent addition to the field of RSV. However, it is considered a promising radionuclide due to its favourable decay properties (17). ^177^Lutetium decays with a physical half-life of 6.73 days. This long half-life is of advantage for shipment to centres far away from the production site. Also, the long half-life leads to prolonged delivery of radiation to the synovium, leading to sustained action. Secondly, it primarily emits beta particles, with an energy of 498 keV (79%) and 177 keV (11%). The beta rays ablate the synovium, thus making it useful for RSV. Further, the beta rays are of low energy, which means that they cause less damage to the surrounding normal tissues. Thirdly, it emits two low-energy gamma photons, 113 keV (6%) and 208 keV (10%), which are useful for imaging. Fourthly, the low energy of beta as well as gamma rays leads to low radiation doses to the hospital staff and the attendants (17, 18).

Hydroxyapatite, with the formula Ca_10_(PO_4_)_6_(OH)_2_, is a natural substance and is a constituent of bone matrix. Hydroxyapatite particles are degraded into Ca^2+^ and PO4^3-^ ions by natural metabolic processes and eliminated from the body over six weeks, thus providing excellent biocompatibility. Thus, hydroxyapatite is one of the most suitable carrier particles for RSV (19).

In the past, RSV with different radioisotopes, and disease states, using different criteria for categorizing responders and non-responders have been published with a wide range of success rates of the procedure (12, 20-25). Several preliminary preclinical studies have explored the use of Lutetium-177 hydroxyapatite as an agent for RSV. These studies demonstrated excellent in-vivo stability of the radioisotope, with complete retention of radioactivity in the joint without evidence of leakage (26, 27). A handful of studies have also been conducted on human subjects which demonstrated the efficacy of Lutetium-177 hydroxyapatite RSV in treating knee joint conditions (28, 29). In light of these findings, we conducted this prospective study to evaluate the efficacy of Lutetium-177 hydroxyapatite RSV for refractory chronic inflammatory arthritis of the knee joint.

## MATERIALS AND METHODS

This prospective study was conducted after receiving ethical clearance from the Institutional Ethics Committee (vide number AIIMS/IEC/2022/3957). The study was registered under the Clinical Trials Registry of India (CTRI) via reference number CTRI/2022/04/041884.

Written informed consent was taken from all patients before enrolling in the study. A total of 24 knee joints in 22 patients were included in the study. Two patients underwent RSV of the same knee twice, and one patient underwent RSV of both knees.

The inclusion criteria was:

1. patients with refractory chronic inflammatory arthritis of the knee joint (>6 months) with evidence of inflammation on bone scan.
2. patients with rheumatoid arthritis unresponsive to conventional medical treatment (NSAIDs, DMARDs, and/or biologicals) for at least 6 months.
3. patients with osteoarthritis unresponsive to conventional medical treatment (NSAIDs) for at least 6 months.

The exclusion criteria included pregnant and breastfeeding women, women planning to get pregnant within the next 6 months, children < 6 years, local skin infection at the injection site, suspected septic arthritis, intra-articular fracture or ruptured popliteal cyst. The exclusion criteria also included severe joint instability due to bone destruction, patients who had undergone arthroscopy or surgery of the affected knee joint within the past 6 week, and patients who gad received RSV for the involved knee joint within the last 6 months.

### Initial assessment

Baseline clinical assessment was done with the following parameters:

i. Knee pain measured by Visual Analogue Scale (VAS), where 0 denoted no pain, and 100 denoted severe or intolerable pain (30).
ii. Tenderness at the knee joint: It was graded on a scale of 0 to 3, wherein 0 indicated no pain on palpation of the knee joint, and 3 indicated withdrawal on palpation of the knee joint (Table 1).
iii. Range of motion at the knee joint: It was graded on a scale of 0 to 3, where 0 indicated maximum range of motion and 3 indicated least possible range of motion (Table 1).
iv. Medication score: It was graded on a scale of 0 to 6, where 0 indicated no medication intake and 6 indicated use of immunosuppressants (Table 1).
v. Oxford knee score (OKS): This is a questionnaire which was used to evaluate patient’s restrictions in performing daily tasks. The total score ranged from 0 to 48. A higher score on the Oxford Knee Score indicated better functional ability, while a lower score indicated more functional impairment in the patient (31-33).
vi. Blood pool index on three phase bone scan: It was calculated according to the method described in Figure 1.

**Table 1:** Baseline clinical assessment parameters.

| Parameter | Score | Interpretation of score |
| --- | --- | --- |
| Pain at the knee joint<br>(according to VAS) | 0 | 0-25 points on VAS |
|  | 1 | 26-50 points on VAS |
|  | 2 | 51-75 points on VAS |
|  | 3 | 76-100 points on VAS |
| Tenderness at the knee joint | 0 | No pain |
|  | 1 | Mild pain |
|  | 2 | Wincing |
|  | 3 | Withdrawal on palpation of joint |
| Range of motion at the knee joint | 0 | >150 degree |
|  | 1 | 100 - 150 degree |
|  | 2 | 50 - 100 degree |
|  | 3 | < 50 degree |
| Medication score | 0 | No medication |
|  | 1 | NSAID one tablet as and when required |
|  | 2 | 1 tablet of NSAID per day |
|  | 3 | 2 - 4 tablets of NSAIDs per day |
|  | 4 | > 4 tablets of NSAIDs per day |
|  | 5 | DMARDs |
|  | 6 | Immunosuppressants |
| Oxford knee score<br>(less score indicates more severity) | 0 | 40-48 points (Satisfactory joint function) |
|  | 1 | 30-39 points (Mild to moderate arthritis) |
|  | 2 | 20-29 points (Moderate to severe arthritis) |
|  | 3 | 0-19 points (Severe arthritis) |

K1 = knee ROI counts in anterior image B1 = background ROI counts in anterior image
K2 = knee ROI counts in posterior image B2 = background ROI counts in posterior image

**Figure 1:**
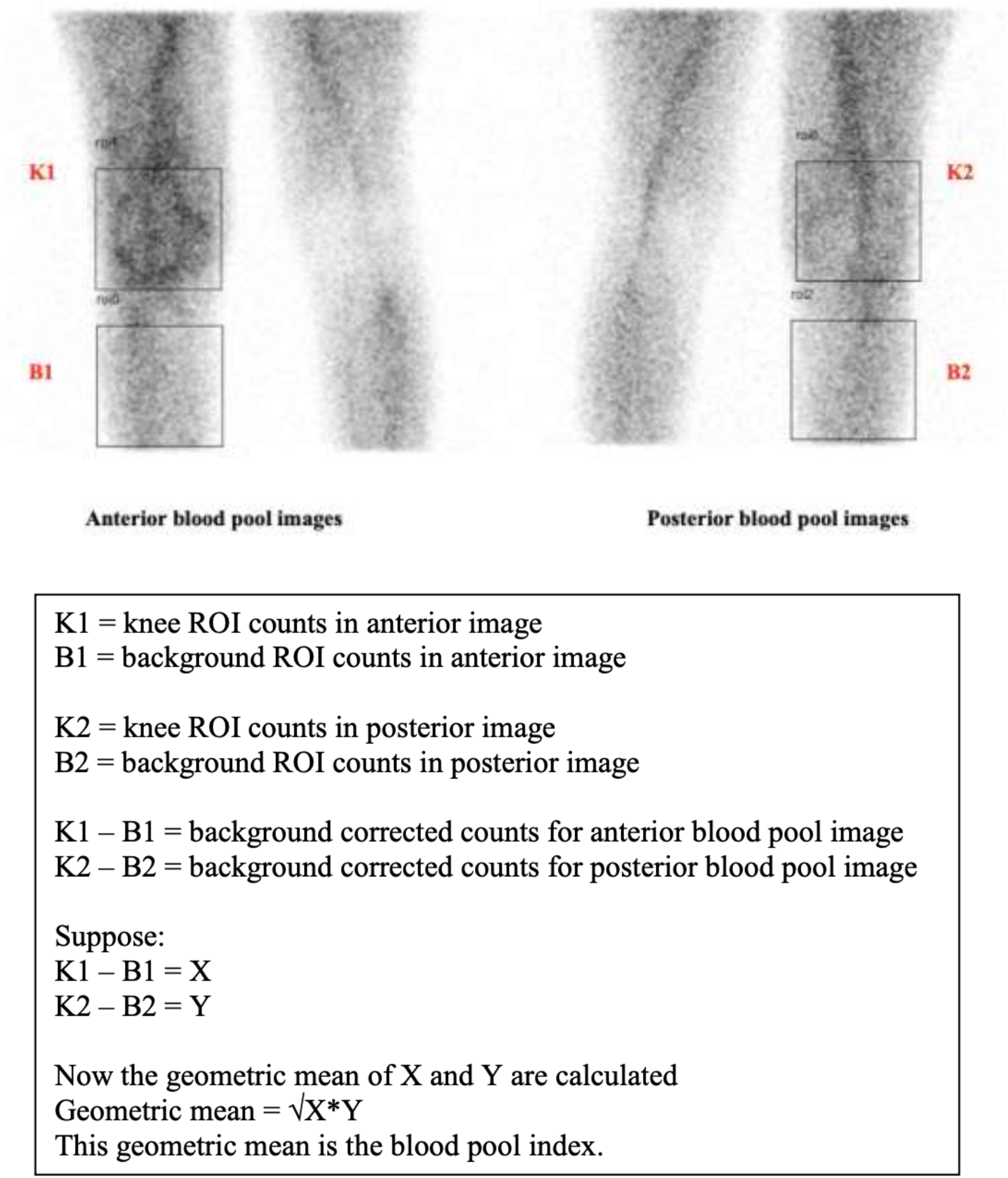
Method used for calculating blood pool index on three phase bone scan

### Technique of radiosynovectomy

The procedure was performed under aseptic conditions on an outpatient basis. The knee joint to be injected was prepped with povidone iodine and alcohol. The knee was then draped with a sterile towel. Local anaesthetic (2% lignocaine) was infiltrated subcutaneously around the injection site. The knee joint was then punctured with a 20-gauge needle via the superolateral approach. 1-2 mL of synovial fluid was aspirated to confirm the intra-articular position of the needle. In case of significant knee joint effusion, the effusion was aspirated first. The syringe containing ^177^Lutetium hydroxyapatite was shaken to disperse the particles evenly. Then 10 mCi of sterile ^177^Lutetium hydroxyapatite was injected into the adult knee through the same intra-articularly placed needle.

The needle through which ^177^Lu-HA was injected was flushed with 0.9% saline. Pressure on the injection site was applied as the joint was put through a mild range of motion to disperse the radionuclide throughout the synovial surface. A small adhesive bandage was applied at the injection site after the procedure. Then a knee brace was applied to immobilize the joint. Radiation safety precautions were followed. The patient was monitored on an outpatient basis for 4 hours for any immediate adverse effects. The knee was supported and non-weight-bearing post-injection. The treated knee joint was immobilized for 48 hours to minimize any leakage of radiotracer from the joint space.

A similar procedure was followed in pediatric age groups with radiopharmaceutical dosage as mentioned below:

Age group: 6 yrs. to <12 yrs. = 3 mCi,

Age group: 12 yrs. to <18 yrs. = 5 mCi

### Post therapy scan

A post-therapy radionuclide scan was taken using a dual head gamma camera with a HEGP collimator within 2 hours of the procedure, using a 20% peak centred over the gamma photopeak of ^177^Lutetium (113 keV and 208 keV) to document intra-articular distribution of the radionuclide and rule out any extravasation.

### Patient follow-up

The patient was reassessed clinically at 4 and 12 weeks after RSV, and a repeat ^99m^Tc-MDP bone scan was performed at 12 weeks after RSV. Finally, the patients were assessed for response assessment criteria as detailed below.

### Statistical analysis

Patients were not excluded from the study on the basis of missing score. Descriptive statistics such as mean and standard deviation (SD) have been used to summarize the baseline clinical and demographic profiles of the patients. For categorical data, non-parametric tests were used; comparison was done using the Freidman test since there were more than two data sets. A post hoc analysis was done using the Wilcoxon sign rank test. For quantitative data that followed a normal distribution, the comparison was done using paired t-test, in case it didn’t follow a normal distribution, a non-parametric Wilcoxon sign-rank test was applied. A p-value of <0.05 was considered statistically significant. Analysis was done by using Statistical Package for Social Sciences (SPSS) software version 26.

### Categorization of responders and non-responders

Patients were characterized into responders and non-responders based on change in percentage of cumulative scores (VAS score, tenderness score, range of motion score and medication score). Responders were defined as patients wherein there was at least 50% decrease in cumulative scores at 3 months as compared to baseline. Non-responders were defined as patients with less than 50% decrease in cumulative scores at 3 months as compared to baseline. Oxford knee score was not taken into account while categorising responders and non-responders, since an increase in OKS indicated favourable outcome, whereas a decrease in rest of the clinical variables indicated better outcome for the patient.

## RESULTS

### Baseline evaluation of the patients

A total of 24 knee joints in 22 patients were included in the study. Two patients underwent RSV of the same knee twice. One patient underwent RSV of both the knees, one followed by the other.

The baseline characteristics of the study population is summarised in Table 2. There were 23 adult and one paediatric patients in the study. The mean duration of disease was 3.56 years. Out of 24 patients, six patients had rheumatoid arthritis, five patients had osteoarthritis, five patients had non-specific inflammatory arthritis, four patients had pigmented villonodular synovitis (PVNS), two patients had inflammatory bowel disease associated arthritis, one patient had juvenile idiopathic arthritis and one patient had prepatellar bursitis.

**Table 2:** Demographic characteristics of the study population.

| Clinical characteristics | Value |
| --- | --- |
| Age (years, mean ± SD) | 46.04 ± 14.55 |
| Age group |  |
| Adult | 23 |
| Paediatric | 1 |
| Duration of disease (years, mean ± SD) | 3.56 ± 4.45 |
| Sex |  |
| Male | 14 |
| Female | 10 |
| Types of arthritis |  |
| Rheumatoid arthritis | 6 |
| Osteoarthritis | 5 |
| Non specific inflammatory arthritis | 5 |
| Pigmented villonodular synovitis | 4 |
| Inflammatory bowel disease associated arthritis | 2 |
| Juvenile idiopathic arthritis | 1 |
| Prepatellar bursitis | 1 |
| Side involved |  |
| Right | 17 |
| Left | 7 |

The baseline clinical scores of the patients are shown in Table 3, and the comparison of the baseline and follow-up clinical scores are displayed in Table 4. As shown in Table 4, there was an improvement in all the clinical parameters (namely pain on the VAS scale, tenderness, range of motion, medication score and Oxford knee score) at 1 month follow-up after RSV, which persisted at 3 months follow-up. Kindly note that a lower score on the Visual Analog Scale (VAS), tenderness, range of motion, and medication scale indicated better patient condition. In contrast, a higher score on the Oxford knee score reflected better patient functioning.

**Table 3:** Baseline clinical scores of the patients.

| **Clinical Scores** | **Values** |
| --- | --- |
| Baseline pain score (0-100) | 67.62 ± 35.93 |
| Score 0 (0-25 points) | 4 |
| Score 1 (26-50 points) | 2 |
| Score 2 (51-75 points) | 4 |
| Score 3 (76-100 points) | 14 |
| Baseline tenderness score (0-3) | 1.91 ± 1.10 |
| Score 0 (No pain) | 4 |
| Score 1 (mild pain) | 3 |
| Score 2 (wincing) | 8 |
| Score 3 (withdrawal on palpation of joint) | 9 |
| Baseline range of motion score (0-3) | 1.29 ± 0.75 |
| Score 0 (>150 degree) | 3 |
| Score 1 (100 - 150 degree) | 12 |
| Score 2 (50 - 100 degree) | 8 |
| Score 3 (< 50 degree) | 1 |
| Baseline medication score (0-6) | 3.00 ± 1.95 |
| Score 0 (No medication) | 1 |
| Score 1 (NSAID one tablet SOS) | 6 |
| Score 2 (1 tablet of NSAID/ day) | 5 |
| Score 3 (2 - 4 tablets of NSAIDs/ day) | 3 |
| Score 4 (2 - 4 tablets of NSAIDs/ day) | 2 |
| Score 5 (DMARDs) | 3 |
| Score 6 (Immunosuppressants) | 4 |
| Baseline Oxford knee score (0-48) | 23.66 ± 7.23 |
| Score 0 (Satisfactory joint function) | 0 |
| Score 1 (Mild to moderate arthritis) | 5 |
| Score 2 (Moderate to severe arthritis) | 14 |
| Score 3 (Severe arthritis) | 5 |

**Table 4:** Comparison of baseline and follow-up clinical scores of the patients.

|  | **Baseline** | **1 month follow-up** | **3 months follow-up** |
| --- | --- | --- | --- |
|  | **Mean ± SD** | **Mean ± SD** | **Mean ± SD** |
| **Pain on VAS (0-100)** | 67.08 ± 35.93 | 36.66 ± 28.07 | 27.70 ± 21.96 |
| **Tenderness (0-3)** | 1.91 ± 1.10 | 0.87 ± 0.89 | 0.54 ± 0.65 |
| **Range of motion (0-3)** | 1.29 ± 0.75 | 0.62 ± 0.64 | 0.45 ± 0.65 |
| **Medication score (0-6)** | 3.00 ± 1.95 | 1.62 ± 1.95 | 1.50 ± 1.81 |
| **Oxford knee score (0-48)** | 23.66 ± 7.23 | 26.66 ± 7.44 | 30.12 ± 7.07 |

We had five clinical variables in our study for analysis: (i) pain on the VAS scale, (ii) tenderness, (iii) range of motion, (iv) medication score, and (v) Oxford knee score. All five of our variables followed a normal distribution.

Out of these five variables, two of the variables, namely pain on the VAS scale and Oxford knee score, were continuous variables. Thus, paired samples t-test was used to determine the statistical significance of these two variables at different time points.

Further, the remaining three variables, namely tenderness, range of motion, and medication score, were categorical variables. Thus, a non-parametric test, the Friedman test, was used to determine the statistical significance of the change in these variables. This was followed by a post-hoc test, the Wilcoxon sign rank test, for intergroup comparison. The results of the paired sample t-test and the post-hoc Wilcoxon sign rank test are displayed in Table 5 below. The p-values which are significant (p<0.05) are marked in bold.

**Table 5:**
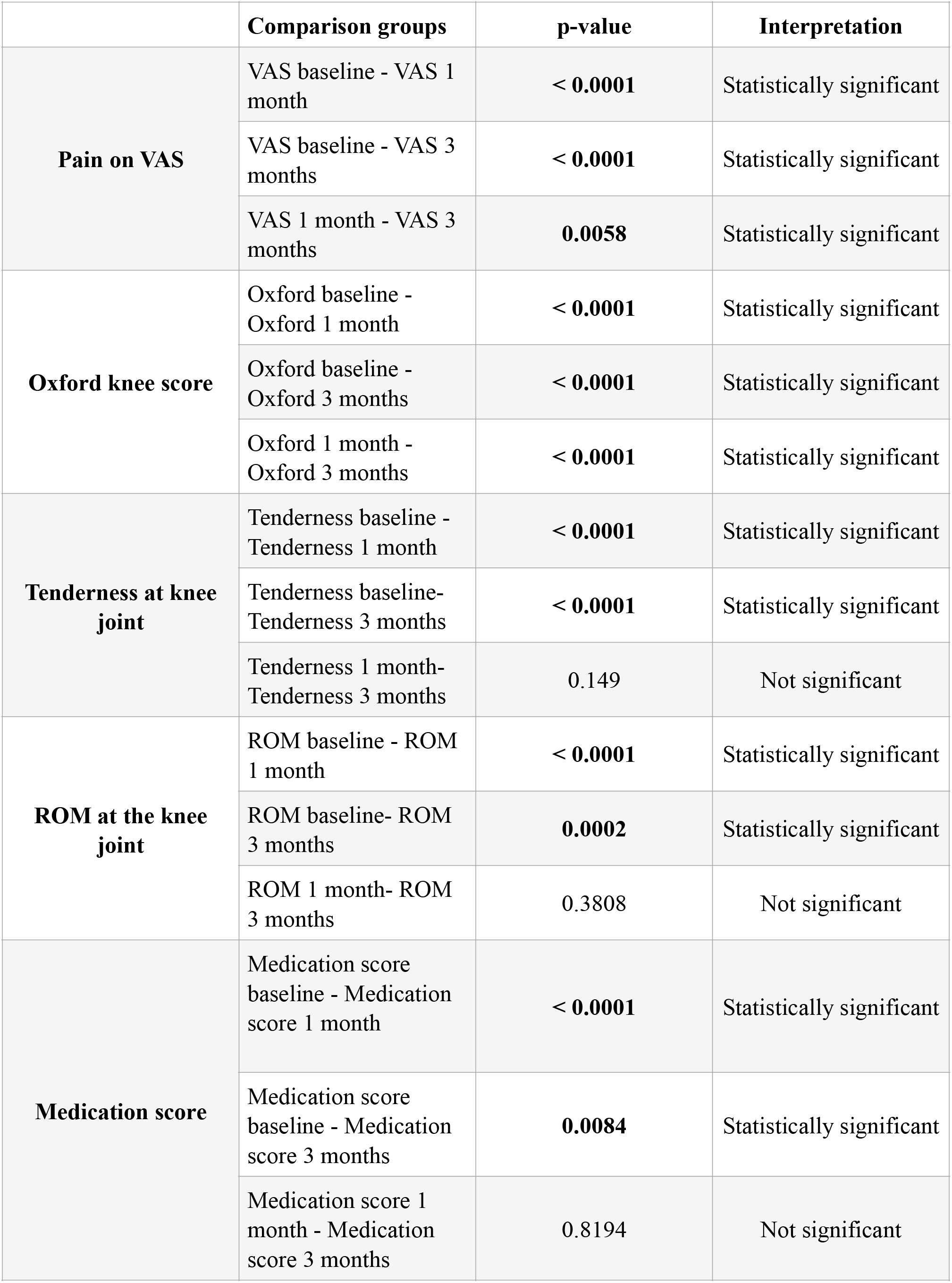
Results of the statistical tests applied to determine statistical significance of the clinical variables at baseline and follow-up.

|  | **Comparison groups** | **p-value** | **Interpretation** |
| --- | --- | --- | --- |
| **Pain on VAS** | VAS baseline - VAS 1 month | **< 0.0001** | Statistically significant |
|  | VAS baseline - VAS 3 months | **< 0.0001** | Statistically significant |
|  | VAS 1 month - VAS 3 months | **0.0058** | Statistically significant |
| **Oxford knee score** | Oxford baseline - Oxford 1 month | **< 0.0001** | Statistically significant |
|  | Oxford baseline - Oxford 3 months | **< 0.0001** | Statistically significant |
|  | Oxford 1 month - Oxford 3 months | **< 0.0001** | Statistically significant |
| **Tenderness at knee joint** | Tenderness baseline - Tenderness 1 month | **< 0.0001** | Statistically significant |
|  | Tenderness baseline- Tenderness 3 months | **< 0.0001** | Statistically significant |
|  | Tenderness 1 month- Tenderness 3 months | 0.149 | Not significant |
| **ROM at the knee joint** | ROM baseline - ROM 1 month | **< 0.0001** | Statistically significant |
|  | ROM baseline- ROM 3 months | **0.0002** | Statistically significant |
|  | ROM 1 month- ROM 3 months | 0.3808 | Not significant |
| **Medication score** | Medication score baseline - Medication score 1 month | **< 0.0001** | Statistically significant |
|  | Medication score baseline - Medication score 3 months | **0.0084** | Statistically significant |
|  | Medication score 1 month - Medication score 3 months | 0.8194 | Not significant |

According to Table 5, there was a significant improvement in all the clinical scores at 1 month compared to baseline. There was also a statistically significant improvement in all the clinical scores at 3 months compared to baseline. For few of the variables, namely pain on VAS score, Oxford knee score and tenderness at the knee joint: there was also a statistically significant improvement at 3 months compared to 1 month. In conclusion, for all the clinical variables, there was an improvement in clinical parameters at 1 month, which persisted till 3 months follow-up.

A visual comparison of the change in clinical scores is depicted in Figure 2 and Figure 3 below.

**Figure 2:**
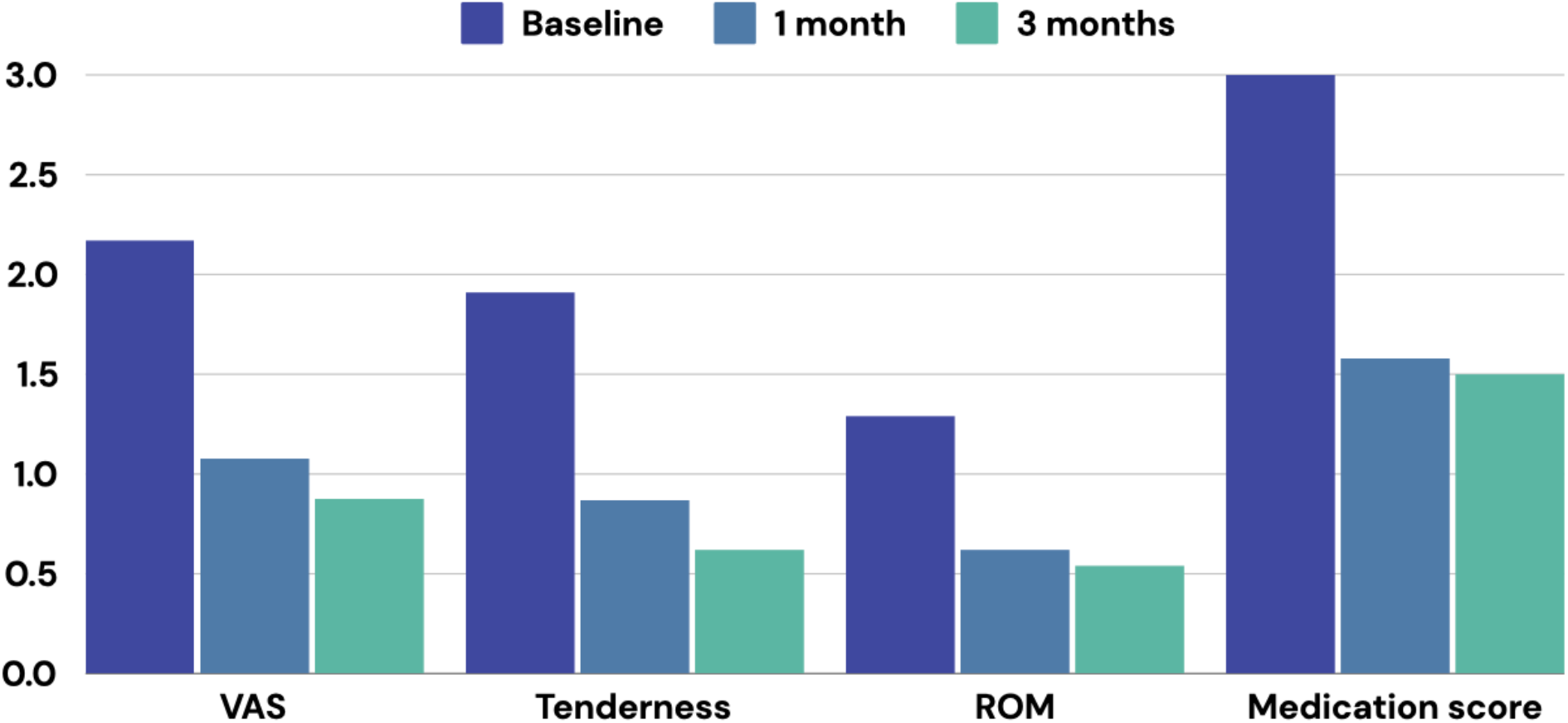
Change in clinical scores at baseline, 1 month follow-up and 3 months follow-up.

**Figure 3:**
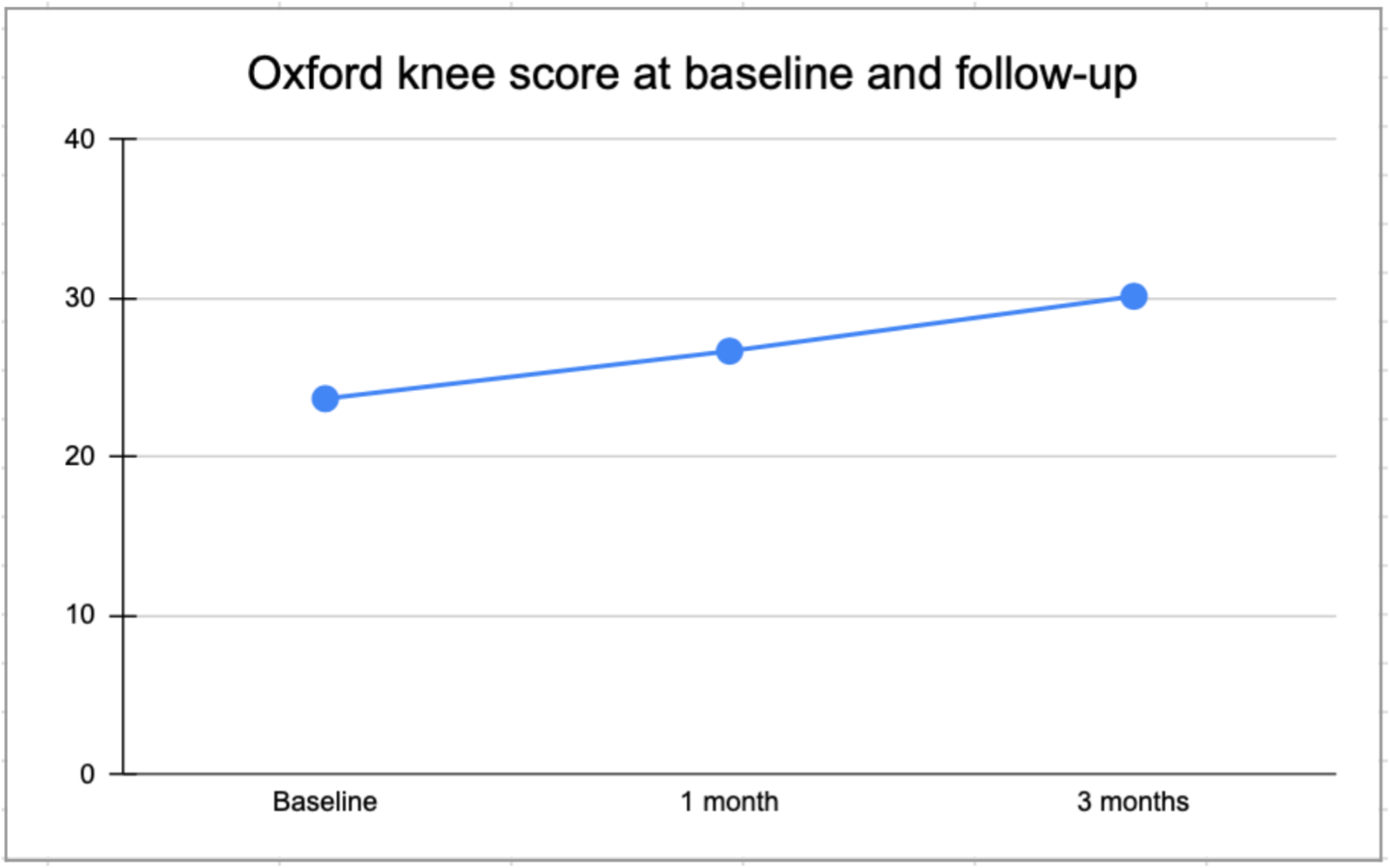
Change in Oxford knee score at baseline, 1 month follow-up and 3 months follow-up.

### Safety

Five out of twenty four patients (20.83%) reported an increase in pain and swelling in the knee joint during the first week following RSV. However, these symptoms resolved in all these patients with the use of analgesics and local ice application. No serious adverse effects were reported in any of the study participants.

Post-therapy whole body scan was taken to document intra-articular distribution of the radiopharmaceutical and to rule out any extravasation. In 23 out of 24 patients (95.83%), there was homogeneous distribution of the radioisotope in the knee joint. However, in one patient (4.16%), during the post-therapy scan, there was a finding of focal increased radiotracer uptake in the left inguinal region. This uptake was on the same side as the side of injection of Lutetium-177 hydroxyapatite. It was initially suspected to be contamination, however, it persisted despite removal of the knee brace, and cleaning the area of skin with soap and water. Thus, it was attributed to uptake in the ipsilateral inguinal lymph node, due to probable extravasation of the radioisotope. The patient was kept on routine follow-up. Dosimetry was not possible as the patient was not willing to come for follow-up whole body scans. The patient improved clinically at 1 month follow up, with sustained improvement till 3 months.

### Categorization of responders and non-responders

Based on the cumulative scores, overall there were 18 responders and 6 non-responders in the study at 3 months after RSV. Thus, the overall response rate of radiosynovectomy was 75%. The plausible reasons for failure of therapy in the six non-responders are listed in Table 6.

**Table 6:** Characteristics of non-responders at 3 months.

| Sl. No. | Diagnosis | Discussion and plausible reason(s) for failure of therapy |
| --- | --- | --- |
| 1 | Prepatellar bursitis | 1. Patient's occupation involved prolonged kneeling, which is a known predisposing factor for prepatellar bursitis (34). Thus despite RSV, continued pressure from kneeling could have counter-acted its therapeutic benefits.<br>2. Insufficient evidence exists in literature for the efficacy of RSV in treatment of prepatellar bursitis (35). |
| 2 | Rheumatoid arthritis | Minimal knee joint effusion, joint space narrowing, reduced joint space, extensive bone destruction. |
| 3 | Pigmented villonodular synovitis | Patient had increased synovial thickness demonstrated on baseline MRI. The maximum range of Lutetium-177 is 1.7 mm (36), thus our radioisotope may not have been able to penetrate the full thickness of the synovium in this patient. |
| 4 | Rheumatoid arthritis | Patient was taking Ayurvedic medications, ? interference by them. |
| 5 | Rheumatoid arthritis | Minimal knee joint effusion, joint space narrowing, reduced joint space, extensive bone destruction. |
| 6 | Osteoarthritis | Grade IV osteoarthritis on X-rays according to Kellgren-Lawrence classification. |

### Disease-specific outcomes after RSV

Overall, there were 24 knees in the study, out of which 18 knees were responders and 6 knees were non-responders. The categorisation of responders and non-responders for each disease subtype is given in Table 7 below, and is pictorially depicted in Figure 4 below.

**Table 7:** Categorization of responders and non-responders for each disease subtype.

| **Disease subtype** | **Total number of patients** | **Number of responders** | **Percentage of responders** |
| --- | --- | --- | --- |
| Rheumatoid arthritis | 6 | 3 | 50% |
| Osteoarthritis | 5 | 4 | 80% |
| Non specific inflammatory arthritis | 5 | 5 | 100% |
| PVNS | 4 | 3 | 75% |
| IBD associated arthritis | 2 | 2 | 100% |
| Juvenile Idiopathic Arthritis | 1 | 1 | 100% |
| Prepatellar bursitis | 1 | 0 | 0% |

**Figure 4:**
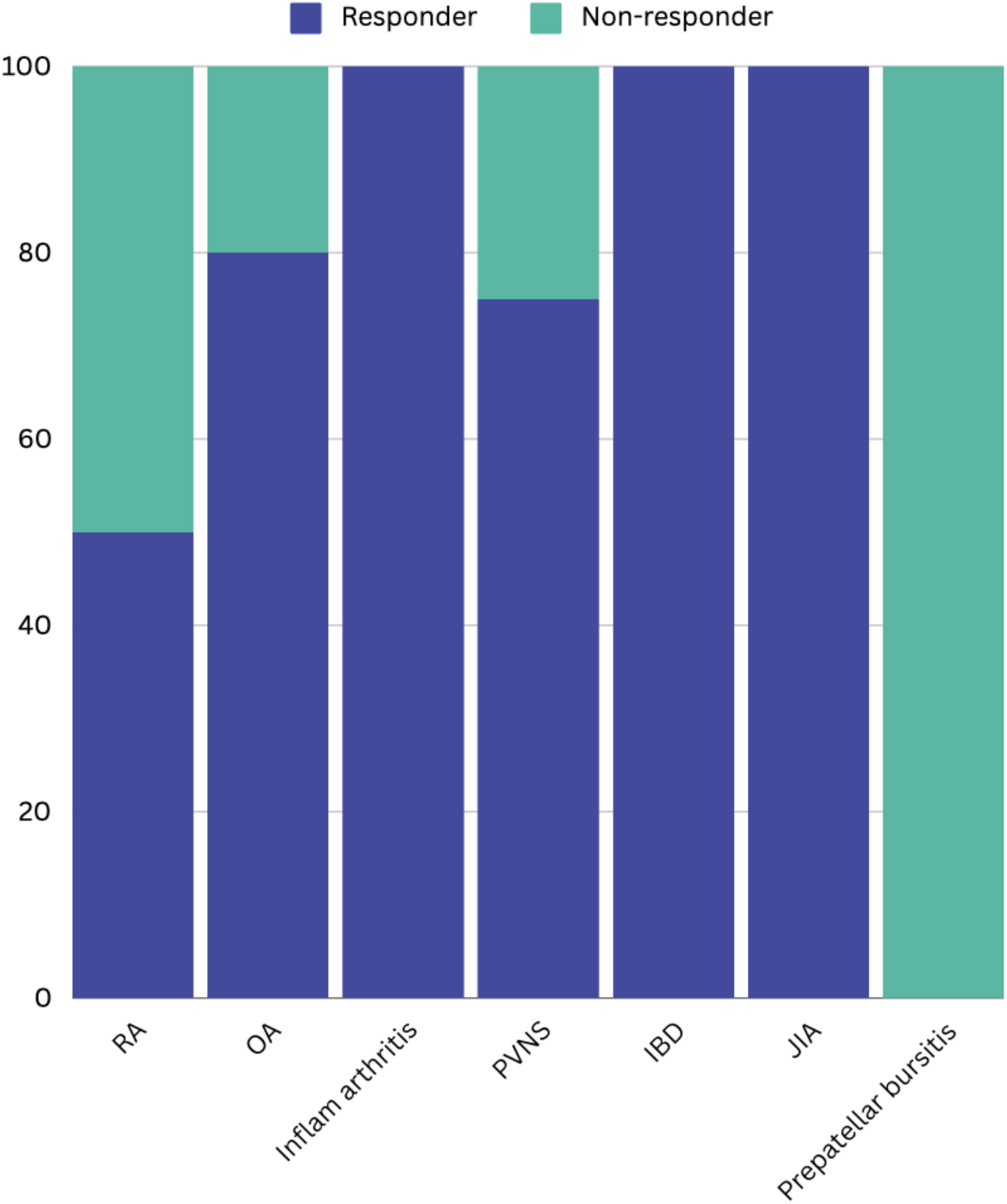
Categorization of responders and non-responders for each disease subtype.

Thus, according to Table 7, the highest response rates were obtained in non-specific inflammatory arthritis, IBD associated arthritis and juvenile idiopathic arthritis. On the other hand, prepatellar bursitis was found to have the lowest response rates amongst the study population.

### Blood pool index on three phase bone scan

In the initial screening phase, all patients underwent a three-phase bone scan. Those exhibiting elevated flow and blood pooling in the affected knee joint on three phase bone scan were selected for RSV. All patients in our study exhibited increased blood pooling in the affected knee on the baseline three phase bone scan. The mean blood pool index at baseline was 12644.23 ± 9673 points (mean ± SD).

The intended follow-up three phase bone scan was originally scheduled for three months post RSV. Unfortunately, due to various issues such as the irregular supply of the radiopharmaceutical, ^99m^Tc MDP, and patient non-compliance, this timing could not be adhered to. Consequently, follow-up bone scans were conducted at different time intervals following RSV. Notably, 9 patients underwent follow up bone scan at three months after RSV.

We restricted our analysis to only those patients who underwent bone scans three months after RSV. Out of the 9 patients who underwent bone scan at three months after RSV, in 6 patients (66.66%), the blood pool index decreased at three months compared to baseline. However, in 3 patients (33.33), the blood pool index increased at three months compared to baseline. A statistical test was not applied to determine if the difference in blood pool index was significant or not, since the data available for follow up blood pool index was small.

The blood pool images of one of these patients at baseline and at 3 months follow-up is shown in Figure 5.

**Figure 5:**
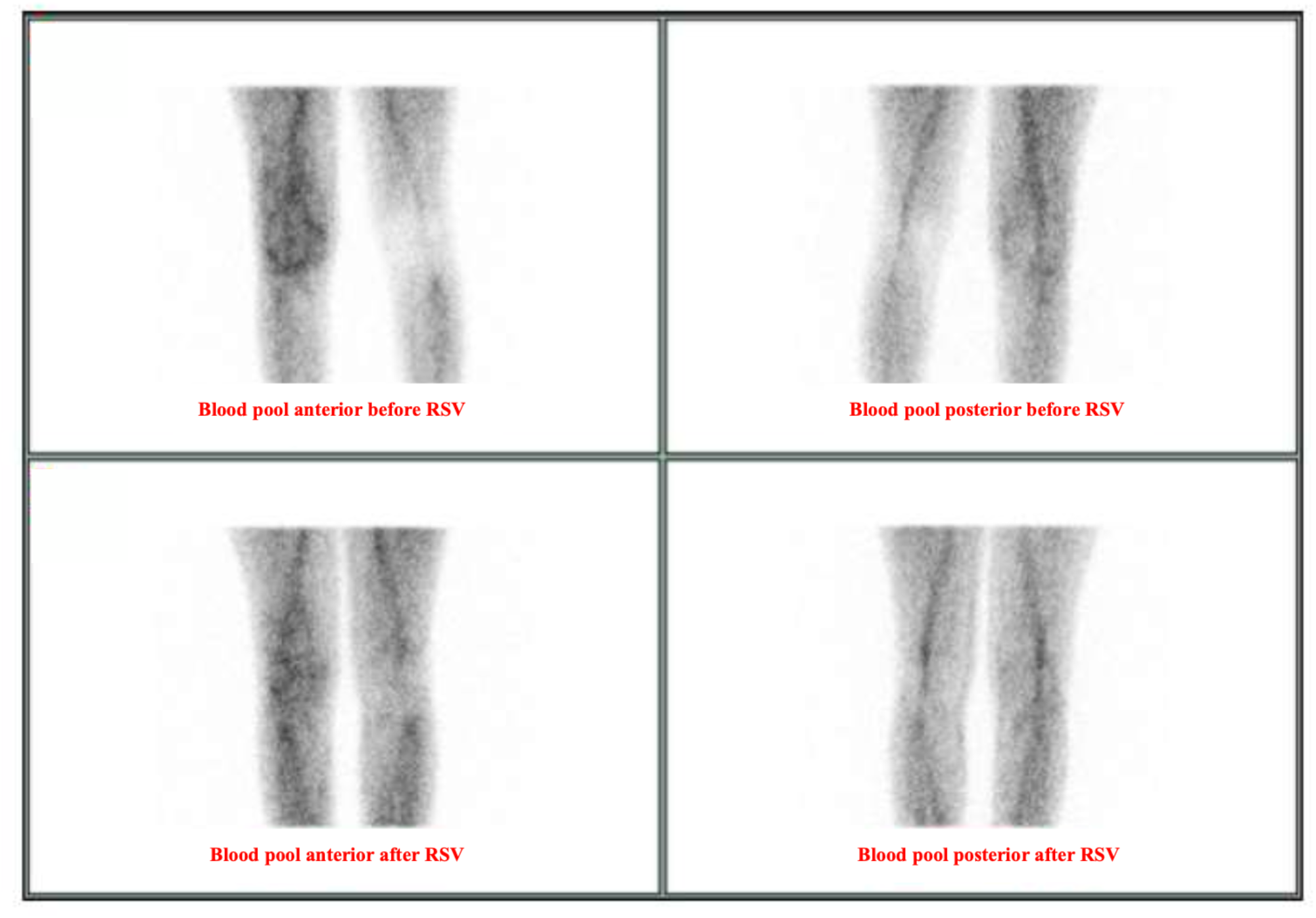
Anterior and posterior blood pool images of a patient with PVNS, before and 3 months after RSV.

## DISCUSSION

In our study, the significant and maintained improvement in synovitis symptoms demonstrate the efficacy of RSV in refractory knee joint inflammatory arthritis. The overall response rate for RSV in our cohort was 75%, closely aligning with findings of a meta-analysis by Kresnik et al., which reported a 72.5% response rate for 2190 treated joints (35). Our results are also consistent with the long term follow-up study by Szentesi et al., wherein excellent and good results were obtained in 71% patients at 5 years, decreasing to 65% at 10 years after RSV (24). Our findings also align with the EANM guidelines, which report a response rate of 60-80% (15).

In our study, 20.83% of the patients reported a transient increase in synovitis, which resolved with local application of ice. There were no major adverse effects reported in the study population. Our findings are consistent with a recent multi-centric study performed by Desaulniers et al., which reported 55 adverse effects out of 392 treated joints, amounting to a 14.03% adverse event rate (38).

Our study also aligns with previous studies that report no long-term increased risk of cancer in patients treated with radiosynovectomy (39, 40).

In our study, the highest efficacy of RSV was obtained in non specific inflammatory arthritis, IBD associated arthritis and JIA patients (response rate of 100%), and lowest efficacy was obtained in rheumatoid arthritis (response rate of 50%). This is in contrary with the study performed by Zalewska et al., who demonstrated highest response rate in rheumatoid arthritis (41). The low efficacy of RSV for patients with rheumatoid arthritis could be explained by the small number of participants in our study.

While RSV has been widely studied globally, Lutetium-177 is a relatively new entrant in the field, resulting in limited research on its use. Kamaleshwaran et al. published a case report on a single patient with elbow joint synovitis secondary to rheumatoid arthritis treated with ^177^Lutetium hydroxyapatite, however, they did not include patient follow-up data (28). A second study by Shinto et al. involved 10 patients with rheumatoid arthritis treated with ^177^Lutetium hydroxyapatite RSV, with a 6 month follow-up (29). However, their sample size was small. A third study by Pragati Jha et al. investigated the usage of ^177^Lutetium tin colloid RSV in 29 patients with rheumatoid arthritis (17). Importantly, all these studies focused solely on patients with rheumatoid arthritis. To the best of the authors’ knowledge, our study is the first in published literature to explore the use of ^177^Lutetium hydroxyapatite RSV in conditions other than rheumatoid arthritis, including osteoarthritis, PVNS, non specific inflammatory arthritis, IBD associated arthritis and prepatellar bursitis.

Similar to our study, many authors including Szentesi et al., Kamaleshwaran et al. and Chrapko et al. have utilized the three phase bone scan to confirm the presence of synovitis before including patients for RSV (24, 28, 42).

Most studies on RSV have employed only clinical and scintigraphic data to evaluate its success (17, 23, 29). Very few studies have incorporated patient-reported outcomes to assess functional status before and after RSV. For example, Amini et al. used the Western Ontario and McMaster Universities Osteoarthritis Index (WOMAC) for patients undergoing ^32^Phosphorus RSV of the knee (21). Prasanna et al, in their study using ^32^Phosphorus for RSV in hemophilic arthropathy, utilized a subjective scoring system, the Tegner Lysholm score, and an objective scoring system, the Modified Knee Society Clinical Rating System (43). In our study, we utilized an objective scale, the Oxford knee score (31-33), since it is easy to use and patient friendly.

Many researchers, such as Shinto et al, have used simultaneous glucocorticoid injection with the radioisotope to reduce the risk of acute radiation-induced synovitis and to avoid skin radiation necrosis (29). During our study, we decided not to administer simultaneous glucocorticoid injection. Steroids can offer short-term symptomatic relief in inflammatory knee arthritis (44), potentially confounding the assessment of efficacy of ^177^Lu-HA. The only exception was in a patient with prepatellar bursitis, for which triamcinolone acetate was injected along with ^177^Lu-HA. This was because the wall lining the prepatellar bursa is thinner compared to the joint capsule, which increases the risk of skin necrosis (35).

In our study, we performed a repeat RSV for one patient who had recurrence of symptoms at four months after the first injection. The interval between the two RSV procedures was 6 months, in accordance with the EANM guidelines (). He responded well to the second injection, and is on routine follow-up. This success of repeat RSV aligns with a study conducted by Stucki et al., wherein they concluded that it was worthwhile to perform repeat RSV in a joint which has initially responded, whereas in a joint that has not responded initially, the chances of success after the second injection are limited (45). Similarly, Vella et al. reported favourable outcomes in 13 rheumatoid arthritis patients who underwent repeat RSV with ^165^Dysprosium (46).

Our study had a few limitations. Firstly, we had a relatively small sample size. Secondly, our follow-up period was for 3 months. Longer follow-ups are needed to assess the long term efficacy of RSV with ^177^Lutetium hydroxyapatite. Thirdly, all patients in our study population did not undergo follow-up bone scan at 3 months after RSV. This was because of non-availability of the radioisotope, ^99m^Tc MDP, at various time points during the study, due to frequent cancellation of the ^99^Molybdenum - ^99m^Technetium generator by the supplier, as well as due to patient non-compliance.

## Supporting information

Oxford Knee Score

## Data Availability

All data produced in the present study are available upon reasonable request to the authors.

