## Supplementary material for "Lutetium-177 hydroxyapatite radiosynovectomy in refractory chronic inflammatory arthritis of the knee joint": Oxford Knee Score

Please answer the following questions. During the past four weeks...

1. How would you describe the pain you usually have in your knee?

- None
- Very mild
- Mild
- Moderate
- Severe

2. Have you had any trouble washing and drying yourself (all over) because of your knee?

- No trouble at all
- Very little trouble
- Moderate trouble
- Extreme difficulty
- Impossible to do

3. Have you had any trouble getting in and out of the car or using public transport because of your knee? (With or without a stick)?

- No trouble at all
- Very little trouble
- Moderate trouble
- Extreme difficulty
- Impossible to do

4. For how long are you able to walk before the pain in your knee becomes seve re? (With or without a stick)?

- No pain >60 minutes
- 16-60 minutes
- 5-15 minutes

- Around the house only
- Not at all – severe on walking

5. After a meal (sat at a table), how painful has it been for you to stand up from a chair because of your knee?

- Not at all painful
- Slightly painful
- Moderately painful
- Very painful
- Unbearable

6. Have you been limping when walking, because of your knee?

- Rarely/never
- Sometimes or just at first
- Often, not just at first
- Most of the time
- All of the time

7. Could you kneel down and get up again afterwards?

- Yes, easily
- With little difficulty
- With moderate difficulty
- With extreme difficulty
- No, impossible

8. Are you troubled by pain in your knee at night in bed?

- Not at all
- Only one or two nights
- Some nights
- Most nights
- Every night

9. How much has pain from your knee interfered with your usual work? (including housework)

- Not at all
- A little bit
- Moderately
- Greatly
- Totally

10. Have you felt that your knee might suddenly give away or let you down?

- Rarely/never
- Sometimes or just at first
- Often, not at first
- Most of the time
- All of the time

11. Could you do household shopping on your own?

- Yes, easily
- With little difficulty

- With moderate difficulty
- With extreme difficulty
- Severe, impossible

12. Could you walk down a flight of stairs?

- Yes, easily
- With little difficulty
- With moderate difficulty
- With extreme difficulty
- Severe, impossible
